# Gaseous micro-embolism (GME) is associated with systemic inflammatory response syndrome (SIRS) after open heart surgery. A missing piece of a complex puzzle?

**DOI:** 10.1101/2022.01.20.22269116

**Authors:** Stefanos Demertzis, Mira Puthettu, Matteo Nafi, Pietro Bagnato, Tiziano Cassina, Stijn Vandenberghe

## Abstract

**Background:** Gaseous micro-embolism (GME) occurring during contemporary open heart surgery is poorly studied. Current understanding of the biological impact of cardiac surgery focuses on the surgical aggression itself together with contact activation of inflammatory cascades by the extracorporeal circulation (ECC), both promoting various degrees of a systemic inflammatory response syndrome (SIRS).

**Methods and Findings:** We prospectively collected data on GME in the ECC circuit according to a quality control protocol during a 12-month period at our institution. Bubbles were measured means of a last generation multi-channel ultrasound measuring unit (BCC300, Gampt GmbH, Meerseburg, Germany) upstream of the arterial line filter. For analysis, bubbles were separated in three size categories: small (S) (10-40 µm), medium (M) (41-200 µm) and large (L) (201-2000 µm). Small bubbles were considered as noise and excluded. A total of 58 out of 70 open heart procedures were included in the final evaluation performed on 58 patients (45 males, 13 females, mean age 66 ± 9 years). Patient baseline data, type of procedure and perfusion data were retrieved. Preoperative treatment with beta-blockers, ACE-inhibitors, calcium-antagonists and statins was considered. Postoperative SIRS was identified according to modified SIRS and qSOFA criteria.

A variably high amount of GME was detected (mean count 847 ± 2560), we focused on M-sized GME (mean count 820 ± 2546, mean volume 233 ± 730 nL). A total of 22 patients (38%) developed SIRS. To account for differences between patient groups (SIRS-/ SIRS+) propensity score (PS) matching was performed on the presence of M-bubbles at or above the 75^th^ percentile (count and volume). The impact of such GME on the development of SIRS was statistically highly significant, as shown by the corresponding average treatment effects (ATE).

**Conclusions:** Significant GME was associated with postoperative SIRS after cardiac surgery in our setting. This novel finding warrants further confirmation.

## Introduction

Gaseous micro-embolism (GME) occurring during contemporary open heart surgery is generally poorly studied and understood. Current understanding sees it as a potential cause of postoperative cognitive decline [1], despite the lack of hard evidence of this association such as a correlated neurological imaging.

Systemic inflammatory response syndrome (SIRS) can complicate open heart surgery. It manifests with hemodynamic instability due to decreased peripheral vascular resistance (so called vasoplegia), fluid accumulation in the third space and increased inflammatory parameters in the blood. Accurate management in the ICU is required to avoid secondary complications.

The use of cardiopulmonary bypass (CPB) or extracorporeal circulation (ECC) is thought to be one of the possible causes of SIRS due to activation of non-specific pro-inflammatory cascades. It is described, that despite the presence of arterial filters with a 40 µm mesh size, air bubbles of larger diameters can be detected in high counts in the arterial line after the arterial filter. Air bubbles embolizing in small arteries and/or capillaries can cause harm by two mechanisms: a reduction in perfusion distal to the obstruction and an inflammatory response[2]. We hypothesized therefore, that GME in the arterial line could be associated with the development of systemic inflammatory response (SIRS) after cardiac surgery.

## Patients and Methods

We prospectively collected data on air-bubble presence in the ECC circuit according to a quality control protocol at our institution. Measurements were carried out during 70 consecutive open-heart procedures. The basic circuit design was identical in all procedures prefabricated customized kits composed of PVC tubing (with heat-exchanger for the cardioplegic solution and suction devices), venous reservoir, oxygenator and arterial filter. De-airing was uniform and according to our standard protocols.

### Bubble measurement

Count and diameter of air bubbles were determined throughout extracorporeal perfusion by means of a last generation multi-channel ultrasound measuring unit (BCC300, Gampt GmbH, Meerseburg, Germany) with sensors placed at three different positions of the extracorporeal circuit. However, for the purpose of this article, measurements are only considered from the probe located directly upstream of the arterial outflow line, thus recording air bubbles going into the patient. The concept and validations of the GAMPT system are described elsewhere [3].

Data were extracted from the BCC300 and synchronized by means of a MatLab algorithm with perfusion log data extracted from the heart-lung machine (Stöckert S5, LivaNova PLC, London, UK). Clinical data were retrieved from the electronic patient record. Data were merged into a spreadsheet table and subsequently anonymized.

### Ethics

All patients signed an informed consent for the use of health-related data for scientific purposes. The study was conducted in accordance with the Declaration of Helsinki and was approved by the Cantonal Ethics Committee (CE TI 4029).

### Bubble data

The measuring range of the bubble counter is 10-2000 µm, while rare overrange bubbles are also counted without dimensional info. Bubble volume is derived automatically by the bubble counter and calculated from the diameters, while also tracking the total accumulated air volume. Data is recorded every second and can be exported as histogram with 10µm bins and as a time line of detected counts and volumes. For analysis, bubbles were separated in three size categories: small-(S) (10-40 µm), medium-(M) (41-200 µm) and large-(L) (201-2000 µm). Small bubbles were excluded and considered as noise.

### Clinical data

Patient baseline data (age, sex), as well as type of procedure (coronary artery bypass grafting (CABG) or valve / aortic), and perfusion data (perfusion- and cross-clamp time) were retrieved. In addition, preoperative treatment with beta-blocker, ACE-inhibitors, calcium-antagonists and statins was identified. For the postoperative period, routinely measured inflammatory parameters (C-reactive protein (CRP), leucocytes) were retrieved and vasoactive intravenous medication was reviewed to identify maximum dose and duration of noradrenaline-infusion, as well, as the perceived need and administration of steroid boluses and / or vasopressin-infusion.

### Definition of SIRS

We defined SIRS according to two frequently used scores SIRS and qSOFA[4] after adapting them to address the initial period after cardiac surgery (intubated and sedated patient).

We confirmed SIRS when two or more of the following criteria were met:

1. CRP at 0 - 24 - 48 hours upon arrival in ICU in doubling increase
2. Leucocyte count at 24 hours > 12000 /µl (as in SIRS scoring)
3. Noradrenaline peak dose > 6 µg/kg/min and / or need of Noradrenaline for >12 h (as surrogate of qSOFA criterion “systolic blood pressure <100”)
4. Administration of steroid bolus and / or vasopressin (as a further surrogate of qSOFA criterion “systolic blood pressure <100”)

Two independent blinded raters assigned the SIRS positive / negative / doubt status. Cases of doubt or of discrepant assigned status were discussed and analyzed in a consensus round. Where needed, more elements (such as temperature, liquid balance) were retrieved from the electronic patient record and a consensus was achieved.

### Statistical analysis

Surgical procedures were classified as CABG only or procedure with opening of the heart chambers (“other”, with or without CABG). This categoric variable was introduced to capture a potentially different bubble-load between these types of operations. New categorical variables were created to identify bubble load at or above the 75^th^ percentile of total count and total volume, respectively. Another categorical variable identified patients with SIRS. Differences between groups were assessed with the Student’s t-test (normally distributed continuous variables) or with the Wilcoxon rank-sum test (categoric variables) as appropriate. To account for differences between the groups and for potential confounding we applied a propensity score (PS) matching procedure. The propensity of measuring bubble values at and above the 75^th^ percentile was calculated by means of logit regression analysis including all available pre-treatment variables. Patients were matched according to the predicted propensity score (1:1 match). Finally, the average treatment effect (ATE) of the matched cohort on the development of SIRS was calculated. To quantify the robustness of the obtained ATE, a sensitivity analysis was performed using the Stata package “tesensitivity”[5]. Significance level was set at p<0.05. All statistical analysis was performed with STATA (version 17.0, StataCrp LLC, College Station, Texas, USA).

## Results

A total of 58 patients were included in the final analysis, operated upon from September 2020 to August 2021. Out of the 70 measured open-heart procedures, 5 patients were excluded because they entered surgery with an already activated inflammatory status (2 patients with acute / subacute endocarditis, 2 patients with mechanical complications of acute / subacute myocardial infarction, 1 patient with an inflammatory reaction of unknown origin). The residual 7 excluded patients were not considered due to incomplete measurements or clearly demonstrated electrical interferences in the BCC300 recordings. A total of 22 patients (38%) developed SIRS, no patient developed clinically evident neurological symptoms or stroke, all patients survived. Clinical characteristics of the included 58 patients are summarized in Table 1.

**Table 1:**
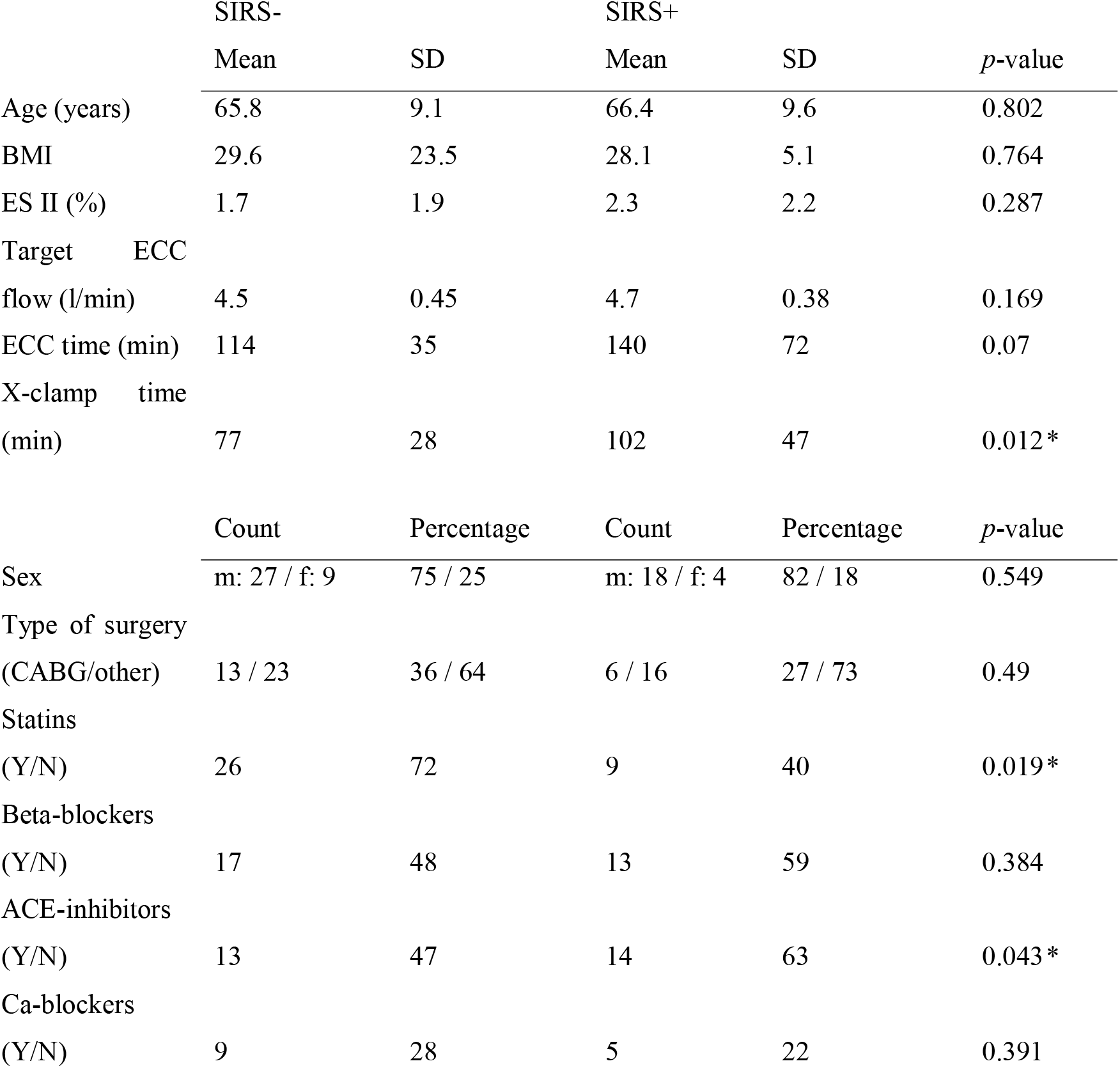
Clinical patient characteristics of the group that did not develop SIRS (SIRS-; n=36) and the group that developed SIRS (SIRS+; n=22) before PS-matching (*: statistically significant differences, SD: standard deviation)

The types of the performed surgical procedures are summarized in Table 2.

**Table 2:** Types of performed surgical procedures (n = 58).

| Type of surgery | SIRS- |  | SIRS+ |  | Total |  |
| --- | --- | --- | --- | --- | --- | --- |
|  | n | % | n | % | n | % |
| CABG | 13 | 36 | 5 | 23 | 18 | 31 |
| CABG & valve(s) | 4 | 11 | 6 | 27 | 10 | 17 |
| Valve(s) | 17 | 47 | 10 | 10 | 27 | 47 |
| Valve(s) & aortic | 2 | 6 | 1 | 1 | 3 | 5 |

Total count and volume of air bubbles was highly variable and with some outliers. Mean total count (S-bubbles (<40µm of diameter) excluded) was 847 ± 25560, mean total volume 233 ± 730 nL.

An overview of the distribution of total, medium-sized (M) and large-sized (L) bubble counts is shown in Figure 1.

**Fig. 1:**
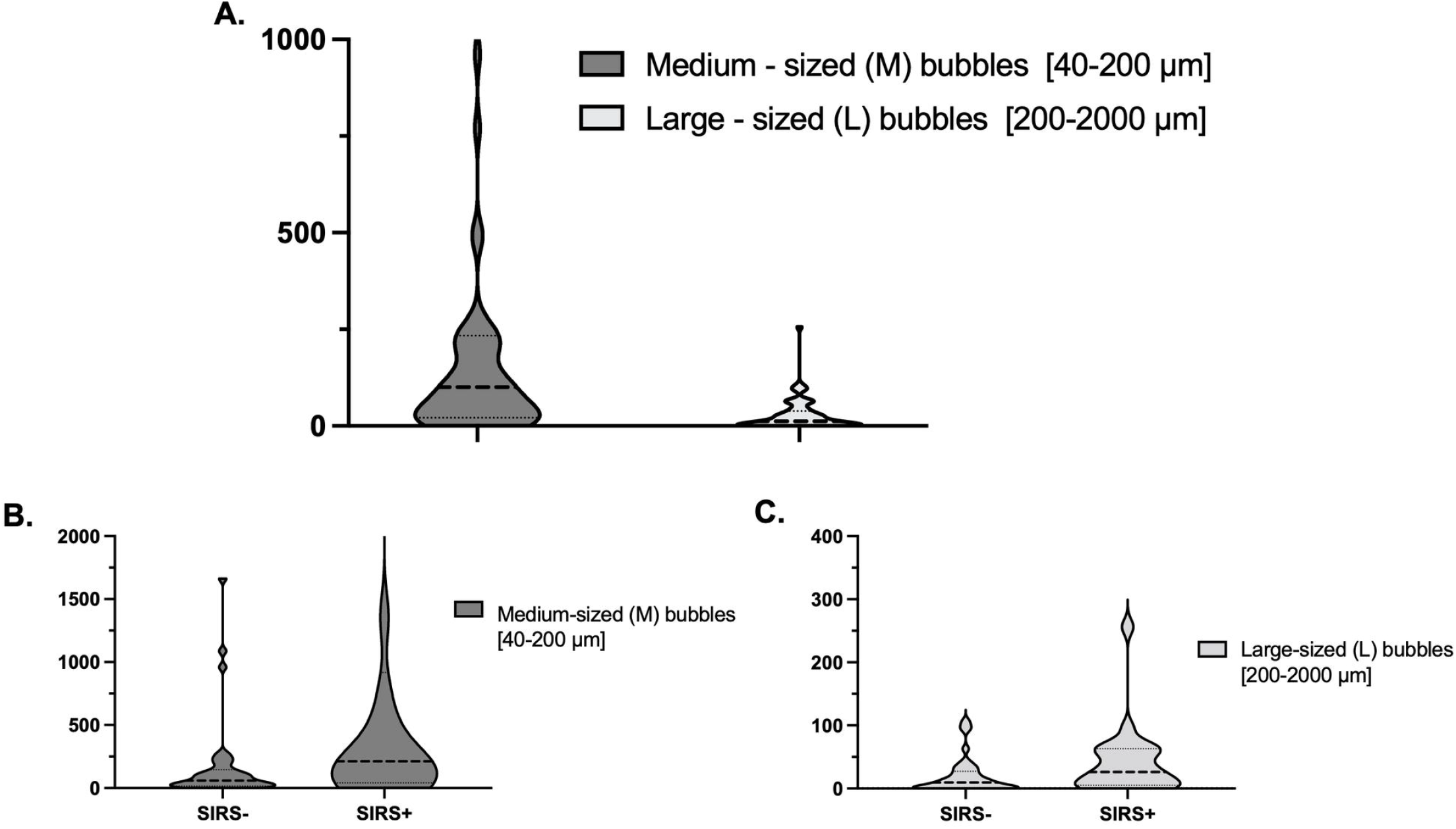
Violin plots (kernel density method) showing the frequency distribution of the measured bubbles A. Counts of all included open-heart procedures, B & C. Counts in patients without and with SIRS in the postoperative period (the dashed lines within the violin represent the median, the dotted ones the quartiles). In A and B the choice of the Y-axis truncated for graphical reasons 4 patients with outlier counts for M-bubbles of respectively: 6008, 6931, 7537 and 16099, which are all included in the analysis. Individual graphs were produced with GraphPad Prism version 9 for Mac, GraphPad Software, San Diego, California USA, www.graphpad.com.

Our analysis focused on the M-sized bubbles due to the high proportion of outliers in the L-sized group. The average values for count and volume for the M-sized bubbles are presented in Table 3. Distribution of the values was not normal, therefore statistical comparison was performed with the Wilcoxon rank-sum test. Patients, who developed SIRS had significantly higher counts and volume of M-sized air bubbles.

**Table 3:**
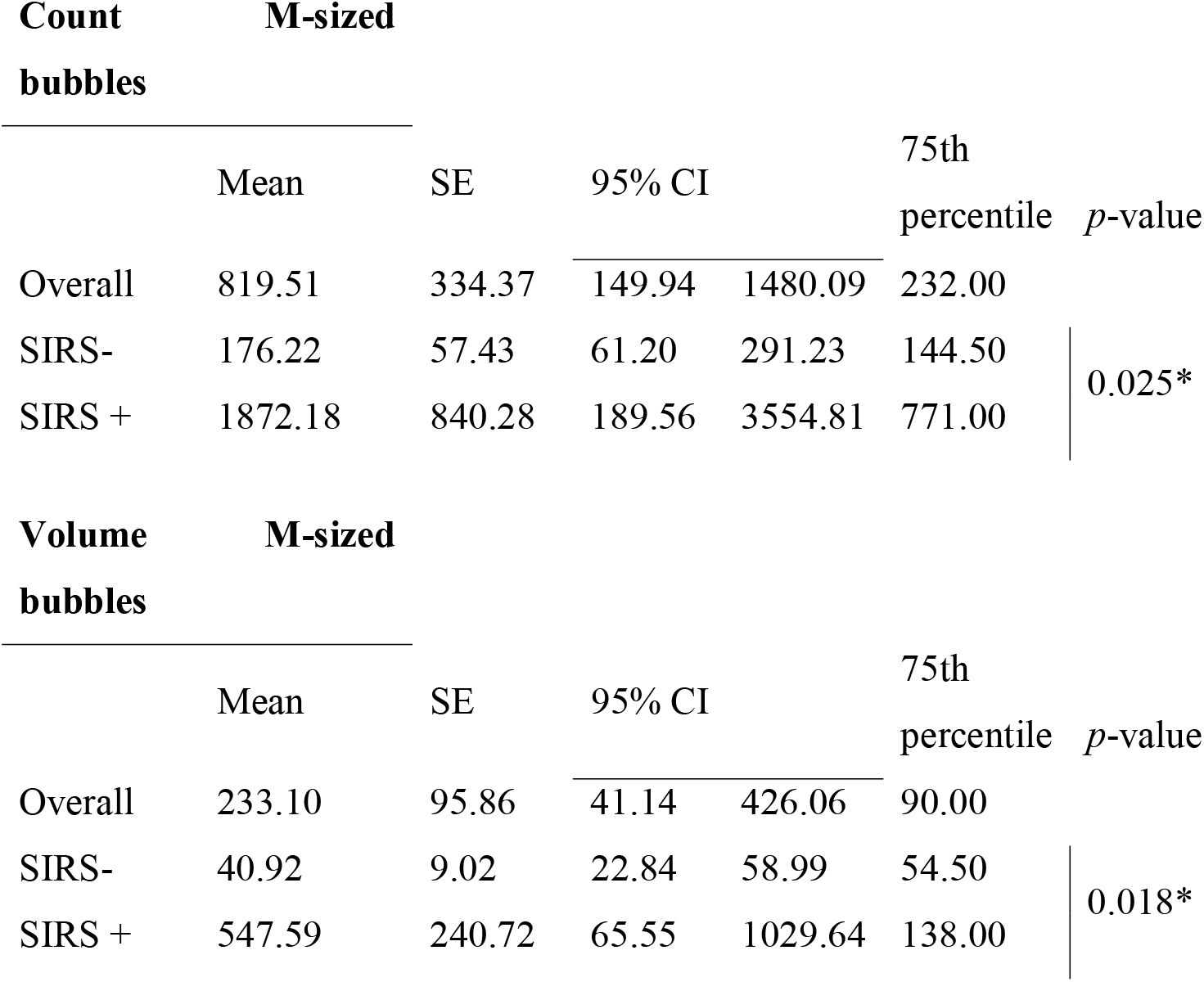
Values for count and volume of the M-medium sized bubbles, overall and divided by groups (*: statistical significance between patient groups).

The categorical variables “CountM_p75” and “VoumeM_p75” were created to depict patients with counts and volume of M-sized bubbles at or above the 75^th^ percentile of the respective distribution. Two PS-matching procedures were performed with distinct “treatment” defined by each one of these variables. The resulting “control” and “treated” groups are shown in Table 4.

**Table 4:**
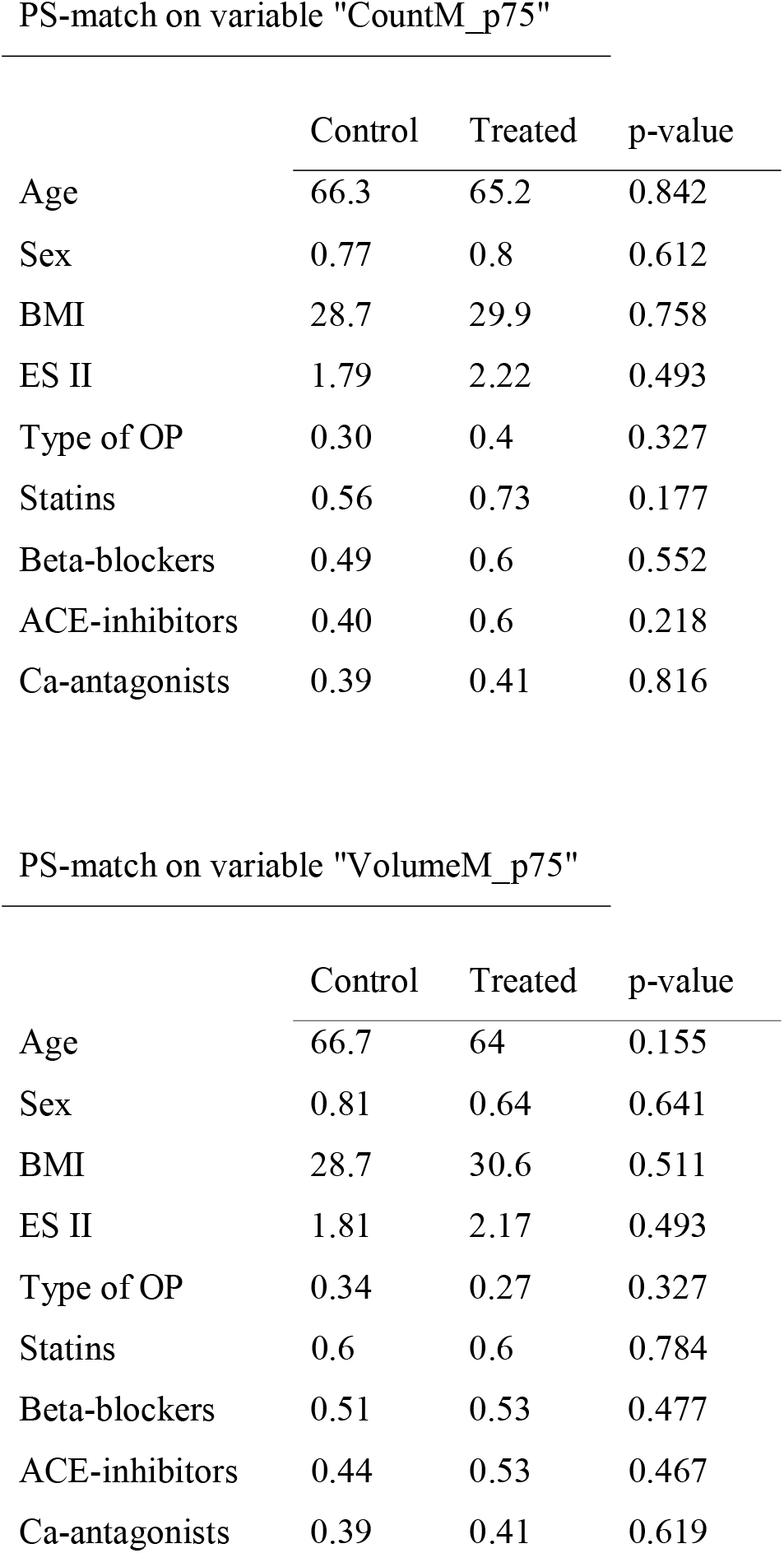
Pre-treatment variables in the “control” and “treated” groups after PS-matching, demonstrating balanced variables between the groups

PS-match on variable "CountM\_p75"
|  | Control | Treated | p-value |
| --- | --- | --- | --- |
| Age | 66.3 | 65.2 | 0.842 |
| Sex | 0.77 | 0.8 | 0.612 |
| BMI | 28.7 | 29.9 | 0.758 |
| ES II | 1.79 | 2.22 | 0.493 |
| Type of OP | 0.30 | 0.4 | 0.327 |
| Statins | 0.56 | 0.73 | 0.177 |
| Beta-blockers | 0.49 | 0.6 | 0.552 |
| ACE-inhibitors | 0.40 | 0.6 | 0.218 |
| Ca-antagonists | 0.39 | 0.41 | 0.816 |

PS-match on variable "VolumeM\_p75"
|  | Control | Treated | p-value |
| --- | --- | --- | --- |
| Age | 66.7 | 64 | 0.155 |
| Sex | 0.81 | 0.64 | 0.641 |
| BMI | 28.7 | 30.6 | 0.511 |
| ES II | 1.81 | 2.17 | 0.493 |
| Type of OP | 0.34 | 0.27 | 0.327 |
| Statins | 0.6 | 0.6 | 0.784 |
| Beta-blockers | 0.51 | 0.53 | 0.477 |
| ACE-inhibitors | 0.44 | 0.53 | 0.467 |
| Ca-antagonists | 0.39 | 0.41 | 0.619 |

The computed average treatment effects (ATEs) were statistically highly significant, indicating that “treatment” characterized by these two variables (high count or volume of mid-sized bubbles) caused more frequently SIRS: for the variable “CountM_p75” the coefficient was 0.6 (p<0.011, 95% CI: 0.46 – 0.75) and for the variable “VolumeM_p75” the coefficient was 0.53 (p<0.001, 95% CI 0.29 – 0.64). The sensitivity analysis showed moderately robust bounds for the estimated ATEs (for “CountM_p75” breakdown at a scalar sensitivity parameter c = 0.136, 6 iterations and for “VolumeM_p75” breakdown at c = 0.215, 10 iterations).

## Discussion

The reported findings show a statistically significant association of higher amounts of mid-sized GME with the development of SIRS after open heart surgery in our setting and thus support the hypothesis of a negative impact of GME on clinical outcomes. The latter might appear obvious, however, solid evidence is still lacking. Therefore, we consider our findings significant, despite the evident limitations of our study: results are from a single center, observational and retrospective study initiated after the evidence of significant amounts of micro-bubbles passing through the bubble-filters and therefore with limited generalizability and lack of control of unknown confounders (known confounders were controlled by means of propensity score matching).

Cardiac surgery and the use of ECC are known to induce a systemic inflammatory reaction. The term “systemic inflammatory response syndrome” (SIRS) describes an intense nonspecific, generalized inflammatory process, which despite some overlaps goes beyond normal postoperative physiology [6]. Morbidity and mortality are increased in patients developing SIRS after cardiac surgery [7]. The prevalence of SIRS after cardiac surgery varies according to the criteria defined for its diagnosis and is reported between 12% and 59% [7,8]. In our setting, prevalence was 38%.

There is an acknowledged issue in identifying SIRS after open heart surgery [7]: patients are admitted to the ICU intubated, ventilated and sedated, having been exposed to ECC, which is unique to this patient population and is generally considered a causative factor for fast-onset postoperative SIRS. This constellation challenges the typical definition criteria for SIRS [4,9] and might cause selection bias due to misclassification. We adopted the two major collections of definition criteria (SIRS and qSOFA) and adapted them to the specific setting of the immediate and early postoperative period after open heart surgery. Differences in assigning a distinct SIRS status emerged between the two blinded raters in borderline patients. A thorough review of each disputed individual record was done to achieve a consensus. Some pharmacological compounds frequently prescribed in patients with heart disease aimed towards reduction of cardiac afterload, could potentially interfere with postoperative vascular resistance, peripheral tissue perfusion and volume shifts. Therefore, we included presence of such drugs (ß-blockers, ACE-inhibitors, calcium antagonists) as potential confounders for the need of postoperative vasopressor support in the calculations to build the propensity score in our analysis.

Direct contact activation of a non-specific immune reaction following interaction of leucocytes, endothelial cells and foreign surface of the ECC circuit, ischemia-reperfusion injury of the endothelium in general and of specific organs (myocardium, lungs, brain, kidneys) and finally endotoxemia due to splanchnic hypoperfusion and bacterial translocation are suggested as possible causes [8]. In any case the subsequent pathophysiological processes seem to be common to all three mentioned possible causes. Activation of several inflammatory cascades occurs (complement-, kallikrein-, cytokine-, activated-neutrophil-, nitric-oxide-, coagulation- and fibrinolytic systems) together with cellular immune response, most of them vigorously interacting with each other [6,10]. Interestingly, almost all attempts to control these mechanisms either pharmacologically or by optimizing the biocompatibility of the ECC circuits were not as successful as initially hoped in tempering the systemic inflammatory response in a measure to influence clinical outcomes [11–14][12–15]. A neglected aspect of open-heart surgery is gas micro-embolism (GME), especially in the contemporary era of modern extracorporeal equipment with adequate bubble filtering. The main part of this work, i.e. detection and measurement of GME using the last version of a specifically developed accurate ultrasonic bubble counter [16,17], was primarily conceived as a quality control study on the basis of similar evidence in the past [18]. The potential association of GME with SIRS after cardiac surgery emerged during internal brainstorming aimed to understand the significance of our findings. We could confirm GME of variable characteristics in all controlled open-heart operations despite all currently available precautions [19][3]. Interestingly enough, we identified a substantial fraction of mainly mid-sized but also large bubbles (40 - 200µm and 201 - 2000 µm of diameter, respecitvely), dimensions well beyond the mesh size of the arterial filters (40µm). Is the presence of these micro-bubbles due to an intrinsic failure of the arterial filters, due to shape deformation of the passing bubbles, or do bubbles break down and re-aggregate to bigger ones after passing the filter? Specific laboratory studies could shed light into this phenomenon.

Arterial gas embolism even only with micro-bubbles can cause tissue ischemia and provoke an inflammatory response [19,20]. In our PS-matched group of patients we were able to identify a significant effect of GME on the occurrence of SIRS after open heart surgery. We were able to detect statistically significant effects when focusing on the higher counts and more specifically of the M-sized GME (41-200 µm in diameter). We believe, this finding makes sense from a biological perspective. Only higher quantities of embolizing M-sized microbubbles are probably sufficient enough to significantly activate or amplify the inflammatory processes without causing clinically detectable tissue ischemia. The systemic impact of GME could be explained by the additive effect of many diffuse local inflammatory reactions caused or amplified by the high counts of clouds of bubbles. This association of GME with postoperative SIRS is not described in literature and warrants further confirmation studies. Significant GME could be indeed the missing element in the complex mechanisms leading to SIRS after open heart surgery.

Acknowledging the limitations of our work, we believe our findings point towards a new mechanism of SIRS after cardiac surgery and we see them as a founded basis to generate hypotheses for future prospective studies. In any case, further analyses are underway to identify possible remedies to reduce GME in our ECC circuits and thereby potentially reduce its negative biological impact on our patients.

## Data Availability

All data produced in the present study are available upon reasonable request to the authors

## Acknowledgments

The precious support of the teams of cardiac surgeons, perfusionists, anesthesiologists, intensivists and the ICU nursing staff of our institute is deeply and thankfully acknowledged. This study was partially funded by grant FF 20134 from the Swiss Heart Foundation.

